# Detection of Somatic Copy Number Deletion of the *CDKN2A* Gene by Quantitative Multiplex PCR for Clinical Practice

**DOI:** 10.1101/2021.09.16.21263412

**Authors:** Yuan Tian, Jing Zhou, Juanli Qiao, Zhaojun Liu, Liankun Gu, Baozhen Zhang, Youyong Lu, Rui Xing, Dajun Deng

## Abstract

**Background:** While the amplification of oncogenes (such as *EGFR*, c-*ERBB2*, c-*MYC*, and c-*MET*) are increasingly driving decision-making for precise cancer treatments, a feasible method to detect somatic copy number deletion (SCND) of tumor suppressor genes is still absent to date.

**Methods:** The genomic coordinates of gene deletion fragments were analyzed using the Catalogue Of Somatic Mutation In Cancer (COSMIC) datasets. Interstitial base-resolution deletion/fusion coordinates for *CDKN2A* were extracted from published articles and our whole genome sequencing (WGS) datasets. The copy number of the *CDKN2A* gene was measured with a multiplex quantitative PCR assay P16-Light and confirmed with whole genome sequencing (WGS).

**Results:** Estimated common deletion regions (CDRs) were observed in many tumor suppressor genes, such as *ATM, CDKN2A, FAT1, miR31HG, PTEN*, and *RB1*, in the SNP array-based COSMIC datasets. A 5.1-kb base-resolution CDR could be identified in >90% of cancer samples with *CDKN2A* deletion by sequencing. The *CDKN2A* CDR covers exon-2, which is essential for P16^INK4A^ and P14^ARF^ synthesis. Using the true *CDKN2A* CDR as a PCR target, a multiplex quantitative PCR assay P16-Light was programmed to detect *CDKN2A* gene copy number with a lower detection limit of 20%. P16-Light was further confirmed with WGS as the gold standard among cancer tissue samples from 139 patients.

**Conclusion:** CDRs are common in many tumor suppressor genes. The 5.1-kb *CDKN2A* CDR was found in >90% of cancers containing *CDKN2A* deletion. The *CDKN2A* CDR was used as a potential target for developing the P16-Light assay to detect *CDKN2A* SCND and amplification for routine clinical practices.

## Background

Somatic copy number variations (SCNVs) of tumor-related genes are landmarks of human cancers [1,2]. Somatic copy number deletion (SCND) and amplification are two kinds of well-known SCNVs. However, current gene copy number detection methods, including microsatellite instability (MSI), loss/gain of heterozygosity (LOH/GOH), fluorescence-in situ hybridization (FISH), whole genome sequencing (WGS) or whole exome sequencing (WES), are not sensitive enough or too costly for routine clinical use. While the amplification of oncogenes (such as *EGFR*, c-*ERBB2*, c-*MYC*, and c-*MET*) is increasingly driving decision-making for precise cancer treatments, clinical applications of SCND of tumor suppressor genes, including *CDKN2A*, are still rare owing to the lack of a feasible detection assay.

The frequency of *CDKN2A* SCND detected by single nucleotide polymorphism (SNP) microarray, WGS or WES was found to range from 30% to 60% in bladder cancer, melanoma, head and neck cancer, pleural mesothelioma, glioblastoma, and esophageal squamous cell cancer (ESCC), with an average frequency of 13% in pan-cancer datasets in The Cancer Genome Atlas (TCGA) (Figure S1A) [2-6]. *CDKN2A* deep deletion is associated with downregulation of *CDKN2A* gene expression, while *CDKN2A* amplification is associated with upregulation of *CDKN2A* gene expression in Pan-TCGA cancers (Figure S1B). It is well known that genetic *CDKN2A* inactivation contributes to malignant transformation, cancer metastasis, and therapeutic sensitivity of cancers to drugs, including CDK4/6 inhibitors and their combination with PD-1 blockade [7-11]. Therefore, a convenient and sensitive assay to detect *CDKN2A* SCND is eagerly awaited.

In the present study, we characterized patterns of estimated genomic coordinates for SCNDs in a set of tumor suppressor genes using the public Catalogue Of Somatic Mutations In Cancer (COSMIC) SCNV datasets and found common deletion regions (CDRs) in many frequently deleted genes. Then, we further defined a 5.1-kb base-resolution CDR within the *CDKN2A* gene using sequencing data for the first time. A sensitive P16-Light assay targeting the *CDKN2A* CDR was established for clinical practice.

## Materials and methods

### COSMIC and TCGA SCNV datasets

SNP6 array-based estimated genomic coordinates of interstitial copy number deletion/fusion of the *CDKN2A* gene in cancer cell lines (n=273) with homozygous *CDKN2A* deletion and estimated genomic coordinates of deep-deleted fragments of *CDKN2A, PTEN, RB1*, and other frequently deleted genes in cancer tissues were downloaded from the Copy Number Analysis (CONA) datasets in the COSMIC project (Data file 1-11) [12].

### Patients, tissues, and DNA preparation

Frozen fresh GC and paired surgical margin (SM) tissue samples were collected from 156 patients in the WGS study [13]. These samples were frozen in liquid nitrogen approximately 30 min after surgical dissection and then stored in a -80°C freezer for 2-5 yrs. Clinicopathological information was also obtained. The 2010 UICC tumor-node-metastasis (TNM) system was used to classify these GCs [14]. Genomic DNA was extracted from these samples with a phenol/chloroform method coupled with RNase treatment. Concentrations of these DNA samples were determined with NanoVue Plus (Biochrom LTD, Cambridge, UK). DNA samples with OD_260nm_/OD_280nm_ ratios ranging from 1.7 to 1.9 were used for the detection of gene copy number as described below.

### Optimized quantitative multiplex PCR assay (P16-Light) to detect CDKN2A copy number

Multiplex primer and probe combinations were designed based on the best multiplex primer probe scores for conserved sequences within the CDR in the *CDKN2A* (HGNC: 1787) and *GAPDH* (HGNC: 4141) gene sequences by Bacon Designer 8 software. Multiplex PCR assays were established according to the Applied Biosystems (ABI) TaqMan universal PCR master mix manual. The performance of these assays for the detection of *CDKN2A* copy numbers was compared with each other. Finally, a multiplex primer and probe combination targeting *CDKN2A* intron-2 was selected (Table 1), and the concentrations of the components were optimized. Each multiplex PCR assay was carried out in a total volume of 20 μL that included 5-10 ng of input DNA, 10 µM of forward and reverse primers and probe for *CDKN2A* intron-2, 10 µM forward and reverse primers and probe for *GAPDH*, and 10 μL of 2x TaqMan Universal Master Mix II with uracil-N-glycosylase (Kit-4440038, ABI, Lithuania). The PCRs were performed in triplicate in a MicroAmp Fast Optical 96-Well Reaction Plate with a barcode (0.1 mL; ABI, China) with an ABI 7500 Fast Real-Time PCR System. The specific conditions of the PCR were as follows: initial incubation for 10 min at 95°C, followed by 40 cycles of 95°C for 20 sec and 58°C for 60 sec. When the Ct value for *GAPDH* input for a sample was 34 or fewer cycles, this sample was considered *CDKN2A* SCNV informative. The specificity of the PCR was monitored through running the gel. Distilled water was used as a no-template control for each experiment.

**Table 1.**
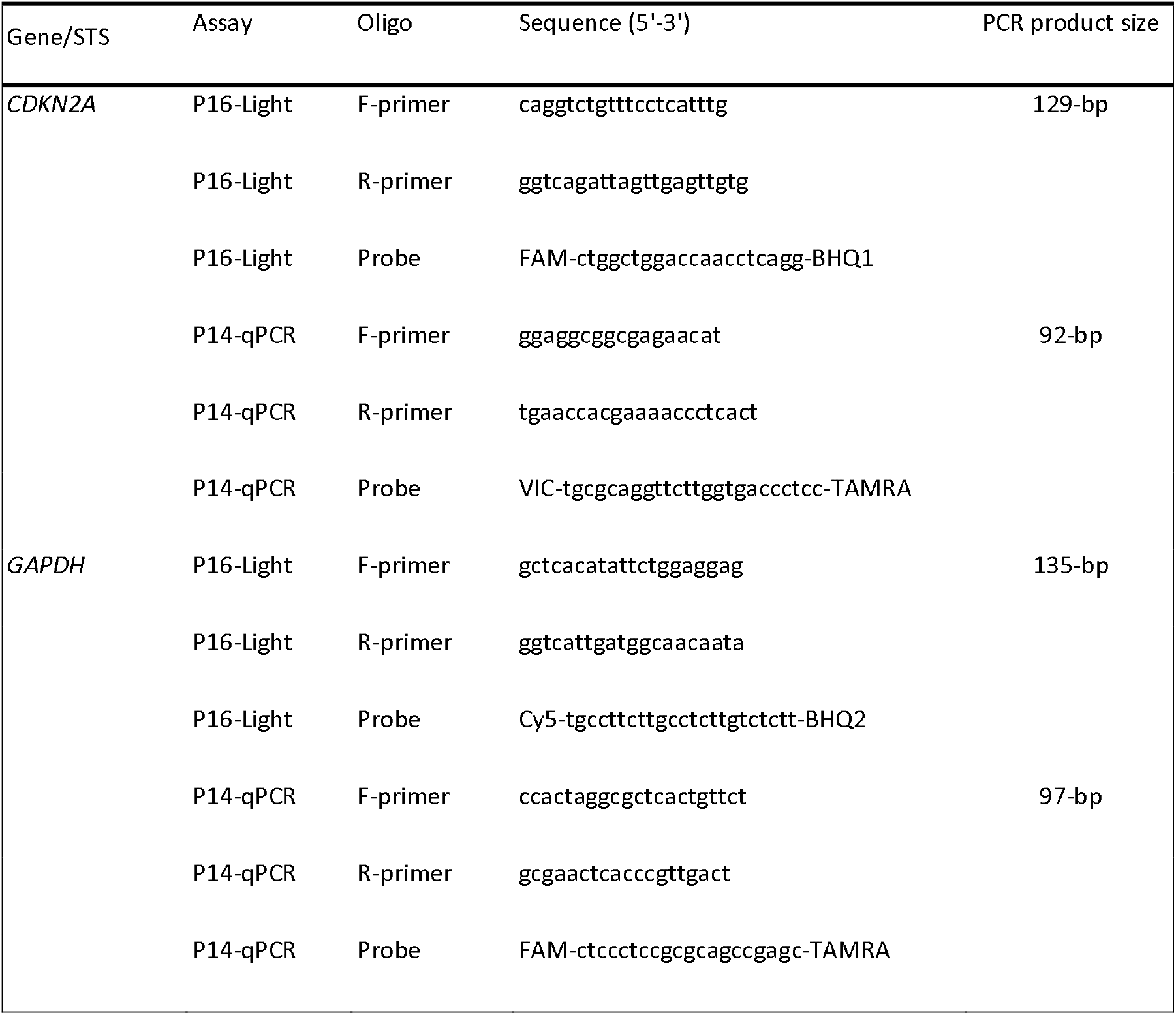
Oligo sequences

### Definitions of CDKN2A CDR deletion positivity and amplification positivity

We used genomic DNA from A549 cells containing no *CDKN2A* allele to dilute genomic DNA from RKO cells containing 2 wild-type *CDKN2A* alleles, and then we set the standard curve according to the relative copy number of the *CDKN2A* gene at different dilutions. The ΔCt value and relative copy number for the *CDKN2A* gene were calculated using the *GAPDH* gene as the internal reference. When the *CDKN2A* copy number in the A549-diluted template was consistently lower than that in the RKO control template and the difference was statistically significant (t test, *p*<0.05), it was judged that the lowest dilution concentration was the detection limit of *CDKN2A* deletion (the difference in *CDKN2A* copy number between the 100% RKO template and 80% RKO template spiked with 20% A549 DNA). When the *CDKN2A* relative copy number in a tissue sample was significantly lower or higher than that of the paired SM sample, the sample was defined as somatic *CDKN2A* CDR deletion-positive or amplification-positive, respectively. For each experiment, the 100% A549, 100% RKO, and 20% A549 + 80% RKO DNA mix controls were analyzed.

### Quantitative detection of CDKN2A/P14^ARF^ exon-1β? copy number by PCR assay (P14-qPCR)

The copy number of *CDKN2A* exon-1β was detected using the primer and probe set (Table 1) as previously reported [15]. When the relative copy number of *CDKN2A* exon-1β in a tissue sample was significantly lower or higher than that of the paired SM sample, the sample was defined as somatic *CDKN2A*/*P14*^*ARF*^ deletion-positive or amplification-positive, respectively.

### Call for *CDKN2A* interstitial deletion/fusion and calculate the purity of cancer cells in the GC WGS datasets

We used Meerkat 23 to predict somatic SVs and their breakpoints in WGS datasets (accession numbers, EGAD00001004811 with 36 × of sequencing depth) for gastric adenocarcinoma samples from 168 patients using the suggested parameters [13]. This method used soft-clipped and split reads to identify candidate breakpoints, and precise breakpoints were refined by local alignments. *CDKN2A* deletion information of 157 GC samples was obtained from WGS datasets. We also estimated copy number profiling over 10-kb windows with Patchwork 28 and calculated the ratio of standardized average depth between normal tissue and tumor tissue (log2R ratio). The purity and ploidy of each tumor were calculated using ABSOLUTE software [16].

### Cell lines and cultures

The *CDKN2A* allele homozygously deleted cell line A549 (kindly provided by Dr. Zhiqian Zhang of Peking University Cancer Hospital and Institute) was grown in RPMI-1640 medium, and the RKO cell line containing two wild-type *CDKN2A* alleles was purchased from American Type Culture Collection and grown in DMEM. The medium was supplemented with 10% (v/v) fetal bovine serum (FBS). These cell lines were tested and authenticated by Beijing JianLian Genes Technology Co., Ltd. before they were used in this study. A Goldeneye™ 20A STR Identifier PCR Amplification kit was used to analyze the STR patterns.

### Statistical analysis

Chi-square or Fisher’s exact tests were used to compare the positive rates of *CDKN2A* SCND or amplification between different groups of tissue samples. Student’s t test was used to compare the proportion of the *CDKN2A* gene copy number between genomic DNA samples. All statistical tests were two-sided, and a *p* value less than 0.05 was considered to be statistically significant.

## Results

### Prevalence of estimated CDRs within various tumor suppressor genes

It has been previously reported that homozygous deletion of approximately 170 kilobase pairs (kb), including the *CDKN2A* locus, can be detected in human cancers by MSI analyses [17]. SCND inactivates the *CDKN2A* gene in 273 human cancer cell lines according to the COSMIC dataset (Data file 1). We found that an 8-kb estimated *CDKN2A* CDR could be detected among these cell lines by ordering “start” genomic coordinates of these breaking points (Figure S2). To investigate the prevalence of CDRs within tumor suppressor genes in human cancer tissues with a high deletion frequency [1,2], we further downloaded the estimated genomic coordinates for deletion fragments that overlapped with these genes. We found that CDRs could be detected not only within the *CDKN2A* gene (Figure 1A; approximately 17 kb) but also within the *ATM* (middle to downstream), *FAT1* (promoter to middle), *miR31HG* (promoter to exon-1), *PTEN* (promoter to exon-1), and *RB1* genes (promoter to intron-2) (Figure 2; approximately 158 kb, 23 kb, 33 kb, 5 kb, and 2442 kb, respectively) (Data files 2-7). No CDR could be observed within *CCSER1, FHIT, LRP1B*, and *WWOX* genes according to the SNP-array data (Data files 8-11).

**Figure 1.**
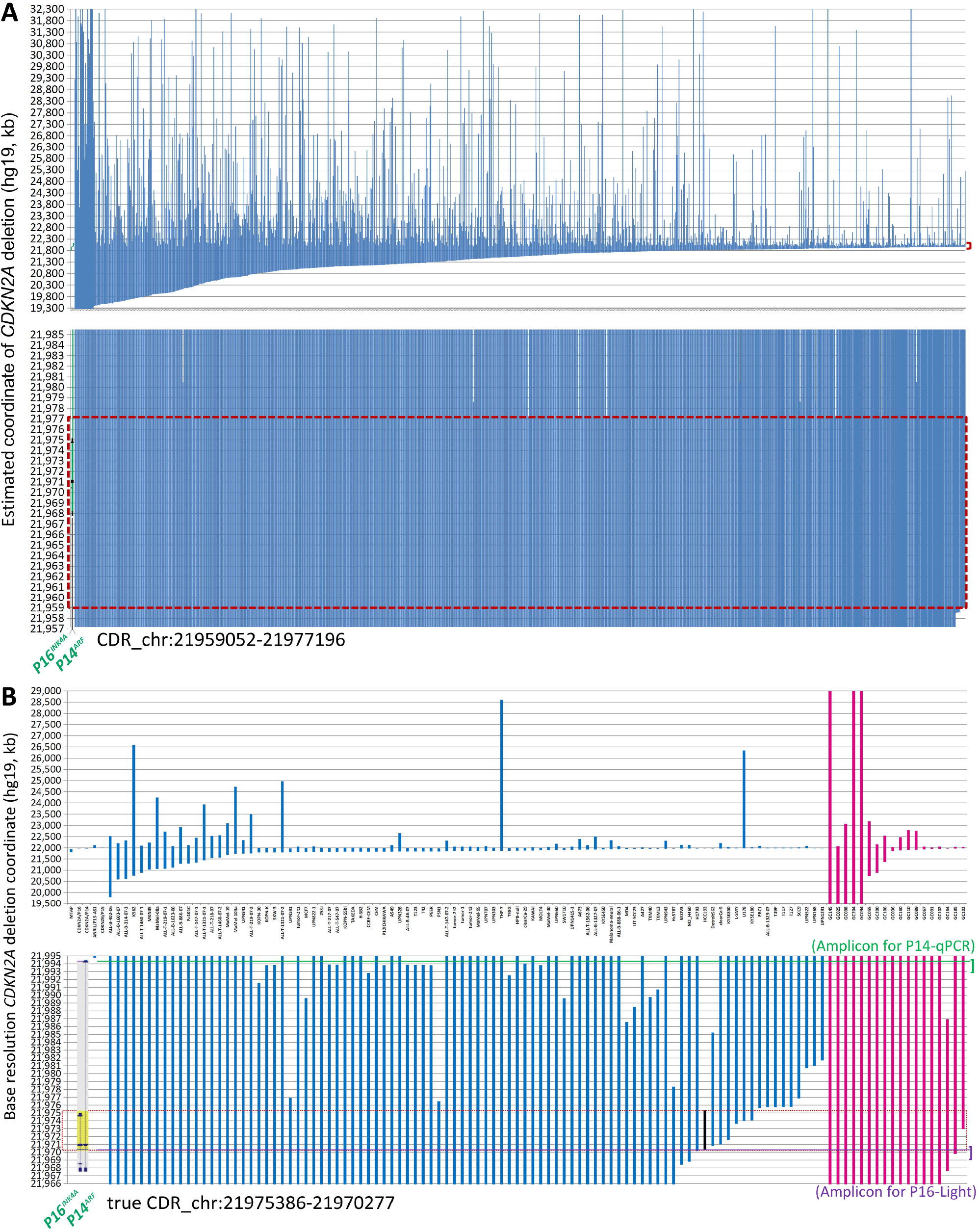
Genomic coordinates of interstitial *CDKN2A* deletion/fusion in human cancer genomes. (**A**) Estimated coordinates of *CDKN2A* deep deletion in cancer tissues according to the COSMIC data. (**B**) True coordinates at the base resolution of *CDKN2A* deletion in cancer cell lines (*n*=92, blue lines) and gastric cancer (*n*=18, purple lines) according to sequencing data. The two top charts display the coordinates of most deletion fragments. The sample ID is labeled under each column. The two bottom charts display the amplified view of these deletion fragments, where the 17-kb and 5.1-kb common deletion regions (CDRs) are highlighted with a red dashed line rectangle. The 5.1-kb true CDR from the *P16*^*INK4A*^ promoter to intron-2 is exactly the same region as the deleted *CDKN2A* fragment in the HCC193 lung cancer cell line (highlighted with a black line). Each line represents a *CDKN2A* deletion fragment. The locations of *P16*^*INK4A*^ and *P14*^*ARF*^ (gray shadow) and exon-1α/1β/2/3 (black dots) are also labeled as landmarks. The positions of amplicons for P16-Light and P14-qPCR are illustrated with violet and green lines, respectively. The detailed deletion coordinates for each sample are listed in Data file 2 and Data file 12.

**Figure 2.**
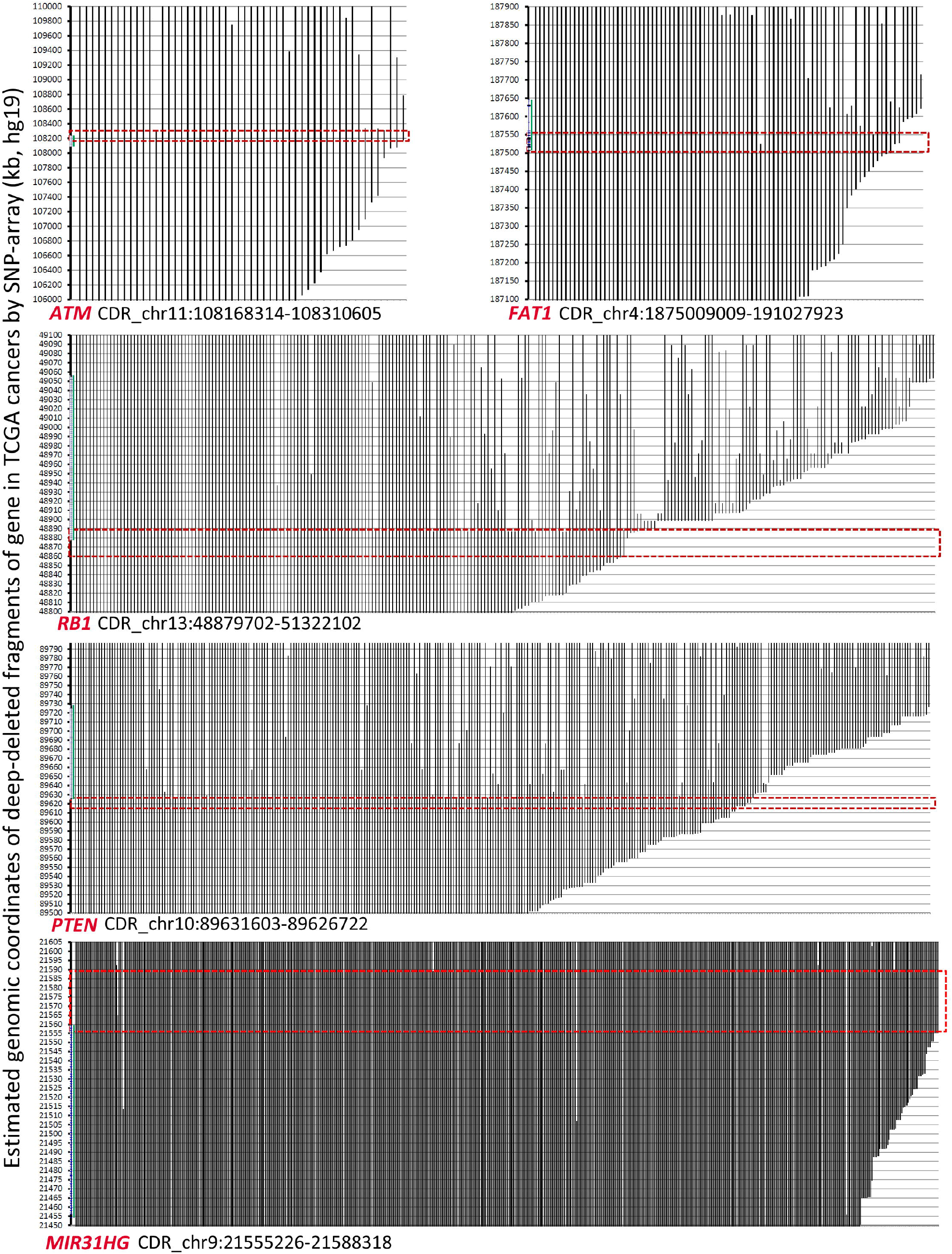
The estimated coordinates of deep-deletion fragments overlapped with tumor suppressor genes *ATM, FAT1, RB1, PTEN*, and *miR31HG* according to the COSMIC data. The common deletion region (CDR) for each gene is highlighted with a red line rectangle. The detailed deletion coordinates for each sample are listed in Data file 3 and Data file 11.

### Characterization of a true CDKN2A CDR at base resolution in human cancers

It was reported that the error in *CDKN2A* breakpoint estimation based on SNP-array data is approximately 10 kb [18]. To characterize the true genomic coordinates of *CDKN2A* deletion fragments in cancers, we extracted base-resolution sequence information of interstitial *CDKN2A* deletions from available published articles and our sequencing data (Data file 12) [19-28]. We found a 5.1-kb CDR (chr9: 21,970,277 - 21,975,386, hg19) that spanned from the *P16*^*INK4A*^ promoter to intron-2 in 83 (90%) of 92 reported cancer cell lines or tissue samples containing interstitial *CDKN2A* deletions (Figure 1B, blue lines). This CDR sequence is the same as the *CDKN2A* deletion fragment in the HCC193 lung cancer cell line [25]. The CDR coordinates were also confirmed in our WGS datasets (average sequencing depth, 36Mendelian Inheritance in Man (OMIM)) of 18 (100%) of 18 GCs [13], in which interstitial *CDKN2A* deletions/fusions were identified (Figure 1B, purple lines; Data file 12).

It is well known that germline *CDKN2A* inactivation can lead to a high predisposition for melanoma and pancreatic cancer [29-31]. Interestingly, we found that 14 (93.3%) of 15 *CDKN2A* allelic variants in the Online Mendelian Inheritance in Man (OMIM) database are located within the CDR sequence, especially in *CDKN2A* exon-2 (Figure S3) [32,33].

In addition, both *P16*^*INK4A*^ and *P14*^*ARF*^ mRNAs are transcribed from the human *CDKN2A* gene at chromosome 9p21 but with different transcription start sites; they share the same exon-2 but have different translation reading frames. Because *CDKN2A* exon-2 located within the true CDR is the essential exon for coding P16^INK4A^ and P14^ARF^ proteins, the above findings indicate that *P16*^*INK4A*^ and *P14*^*ARF*^ are coinactivated in 87% (96/110) of human cancer cell lines and tissues containing *CDKN2A* CDR deletion (Figure 1B).

### Establishment of a convenient PCR assay (P16-Light) to detect somatic CDKN2A CDR deletion

The current clinical method FISH for detecting SCND is composed of a set of biotin-labeled probes that should cover at least 50-kb DNA sequences. Thus, FISH is not a suitable method for detecting the copy number deletion of the 5.1-kb *CDKN2A* CDR. To provide a convenient method for routine clinical use, we designed and experimentally evaluated a set of multiplex quantitative PCR assays and finally optimized the *CDKN2A* CDR-specific quantitative multiplex PCR assay called P16-Light for detecting the copy number of a 129-bp amplicon within the *CDKN2A* intron-2 (Figure 3A), which covers 86% (94/110) of known *CDKN2A* deletion fragments (Figure 1B, violet line).

**Figure 3.**
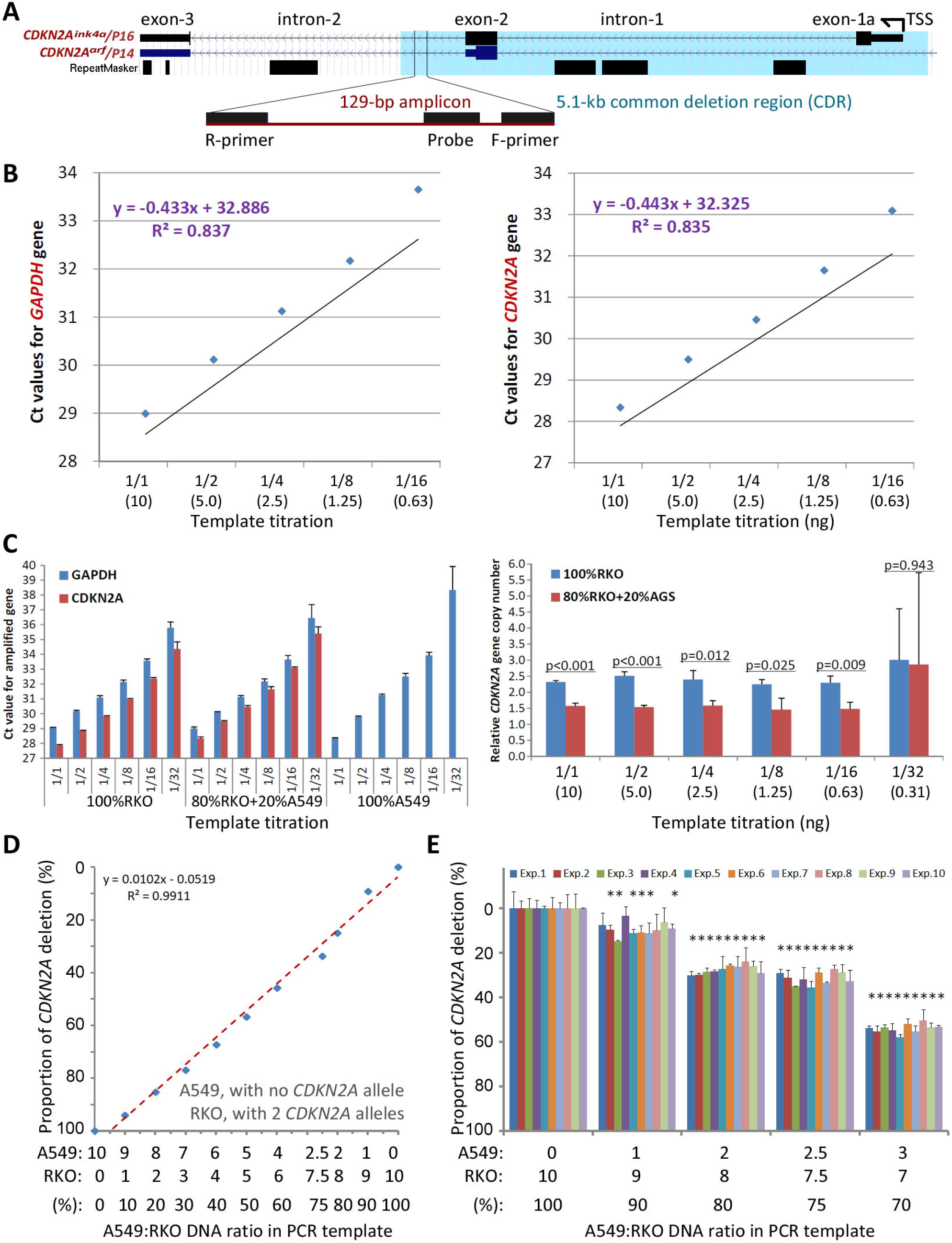
Detection of the copy number of *CDKN2A* intron-2 with quantitative gene-specific multiplex PCR (P16-Light). (**A**) The location of the 129-bp amplicon within the common deletion region (CDR) and its host genes. (**B**) The amplification efficiency of two amplicons for the *GAPDH* and *CDKN2A* genes in the template titration assays using standard DNA samples from RKO cells (with two wild-type *CDKN2A* alleles) and A549 cells (with a homozygous *CDKN2A* deletion). (**C**) Effects of the amount of template DNA on the efficiency of PCR amplification for amplicons in the *CDKN2A* and *GAPDH* genes (left chart) and detection of the relative *CDKN2A* gene copy number (right chart). (**D**) The linear relationship between the proportion of *CDKN2A* copy number deletion and ratios of RKO cells spiked with A549 cells. (**E**) Stability of the proportion of the *CDKN2A* copy number deletion by P16-Light during ten experiments over different days. The RKO cell DNA templates were spiked with 0, 10%, 20%, 25%, and 30% A549 cell DNA. Each column represents the average proportion of *CDKN2A* copy number deletions in triplicate. Exp. 1 - 10: the results of 10 repeated experiments performed on different days. \**P*<0.05.

The copy number of the *GAPDH* gene was used as the internal reference. Genomic DNA from human A549 cells (with homozygous deletion of *CDKN2A* alleles) and RKO cells (with 2 wild-type *CDKN2A* alleles) were used as *CDKN2A* CDR deletion-positive and deletion-negative controls, respectively. The amplification efficiencies of the two amplicons in *GAPDH* and *CDKN2A* were very similar (Figure 3B). No template inhibition was observed when the amount of template DNA ranged from 10 to 0.63 ng (Figure 3C). The proportions of *CDKN2A* CDR copy number were linearly correlated with the ratios (0 - 100%) of RKO cell DNA and A549 cell DNA in the input mixtures (10 ng/reaction) when the A549 DNA was spiked in at different proportions for the P16-Light analyses (Figure 3D). Furthermore, there was a high reproducibility when DNA with homozygous deletion of *CDKN2A* was present in ≥20% of the cells verified in ten experimental repeats performed on different days (Figure 3E). Thus, when the proportion of *CDKN2A* copy number was significantly decreased (or increased) in a sample relative to the paired normal control (t test, *p*<0.05) in the P16-Light analyses, the sample was defined as *CDKN2A* SCND-positive (or amplification-positive).

### Comparison of P16-Light with WGS datasets

As we described above, information on interstitial copy number deletion/fusion of the *CDKN2A* gene was extracted from WGS datasets for 156 of 168 GC patients enrolled in a GC genome study [13], and a total of 18 *CDKN2A* deletion/fusion coordinates at the base resolution were detected in 17 (10.8%) GCs (Data files 12 and 13). To compare the performance of P16-Light with WGS, we analyzed the status of SCNVs, including SCND and amplification, of the *CDKN2A* gene in 156 of these GCs with enough genomic DNA samples with P16-Light using the paired SM sample as the diploid reference (Data file 13). *CDKN2A* SCND and amplification were detected in 40 (25.6%) and 34 (21.8%) of these GCs, respectively. The P16-Light analysis was confirmed by the WGS results: the frequency of *CDKN2A* SCND (or amplification) by P16-Light was significantly higher (or lower) in 17 GCs containing interstitial *CDKN2A* deletion/fusion than in 139 GCs without interstitial *CDKN2A* deletion/fusion (chi-square test, *p*<0.028; Figure 4A). These results also indicate that there is a significantly higher sensitivity for detecting *CDKN2A* SCND by the quantitative P16-Light assay than the hemi-quantitative WGS.

**Figure 4.**
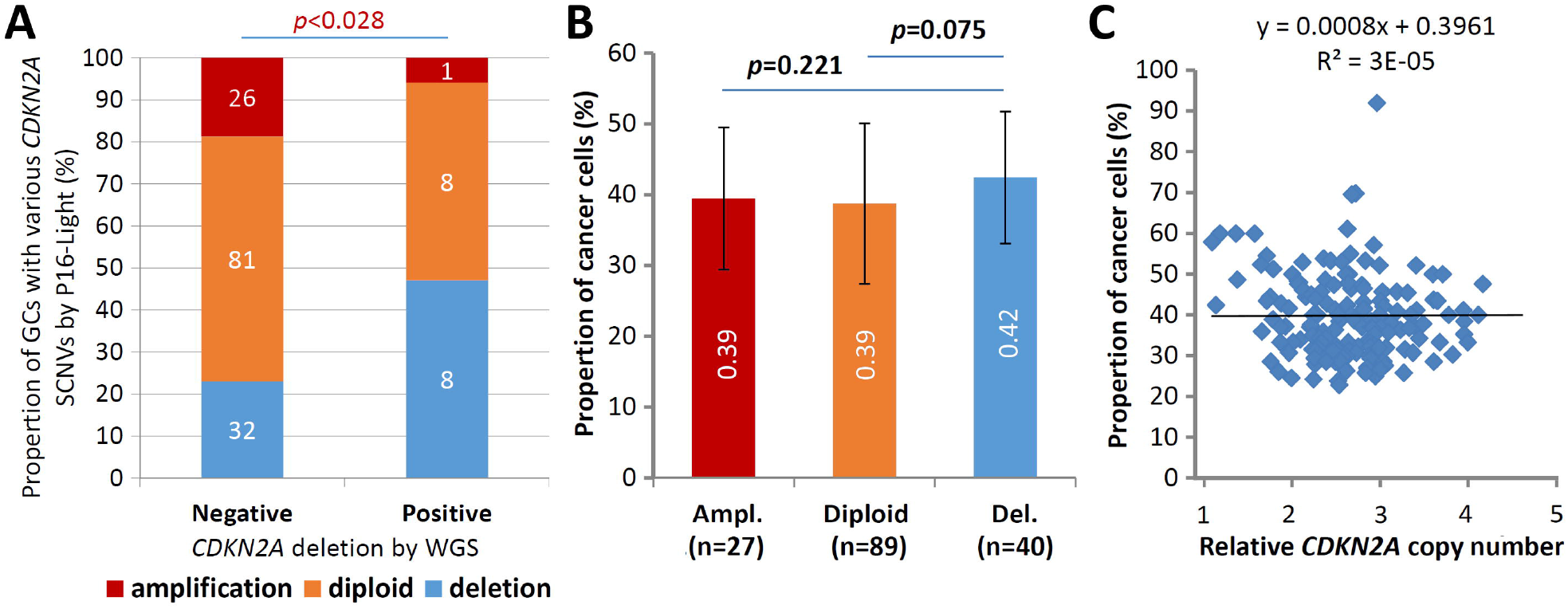
Comparisons of somatic copy number variations (SCNVs) of the *CDKN2A* gene in gastric carcinoma samples (GCs) from 156 patients in the P16-Light and WGS (30Mendelian Inheritance in Man (OMIM)) analyses. (**A**) The states of *CDKN2A* SCNVs by P16-Light (relative to paired surgical margin reference) in GC groups with and without *CDKN2A* deletion/fusion in the WGS analysis. (**B**) Comparison of the proportion of cancer cells (or sample purity; by WGS) in GC groups with various *CDKN2A* SCNVs by P16-Light. The average proportion of cancer cells in each GC group is labeled. (**C**) Correlation analysis between the proportion and relative copy number of the *CDKN2A* gene in GCs.

Moreover, it is well known that the proportion of cancer cells in tissue samples (i.e., sample purity) may affect the detection values of various genome data. To study whether the cancer cell proportion disturbs the detection of *CDKN2A* SCNVs, we calculated the cancer cell proportion in the above GC samples using WGS data (Data file 13). We found that the difference in sample purity between GC subgroups with different CDKN2A SCNV statuses was not statistically significant (t test, *p*=0.075; Figure 4B), although the proportion was slightly higher in GCs with *CDKN2A* SCND than in those without CDKN2A SCND. No correlation was observed between the proportion of cancer cells and the relative copy number of the *CDKN2A* gene among these GCs (Figure 4C).

### Comparison of P16-Light with P14-qPCR assay

The P14-qPCR assay was previously established for detecting the copy number of *CDKN2A/P14*^*ARF*^ exon-1β [15]. Two amplicons in the P16-Light and P14-qPCR assays cover 98% (108/110) of known *CDKN2A* deletion fragments (Figure 1B, violet and green lines). Therefore, we further compared the performance of P16-Light, P14-qPCR, and their combination using GC and paired SM samples from patients who were recently included in the cross-sectional cohort in our association study [34]. GC samples (*n=*139) with enough genomic DNA were used in P14-qPCR analysis (Data file 14). The SCND-positive rate for *P14*^*ARF*^ was similar to that for the *CDKN2A* CDR (31.7% *vs*. 36.7%) (Table 2). *CDKN2A* SCND was found only in 19 GCs by both assays. While *CDKN2A* CDR SCND by P16-Light was significantly associated with distant metastasis of GC (odds ratio=4.09, *p*<0.001), no association was observed between GC metastasis and *P14*^*ARF*^ SCND by P14-qPCR. Using merged *CDKN2A* SCND data (*CDKN2A* CDR SCND-positive and/or *P14*^*ARF*^ SCND-positive), only a weaker association was observed. These results suggest that individual P16-Light alone may be good enough for detecting *CDKN2A* SCND in tissue samples.

**Table 2.**
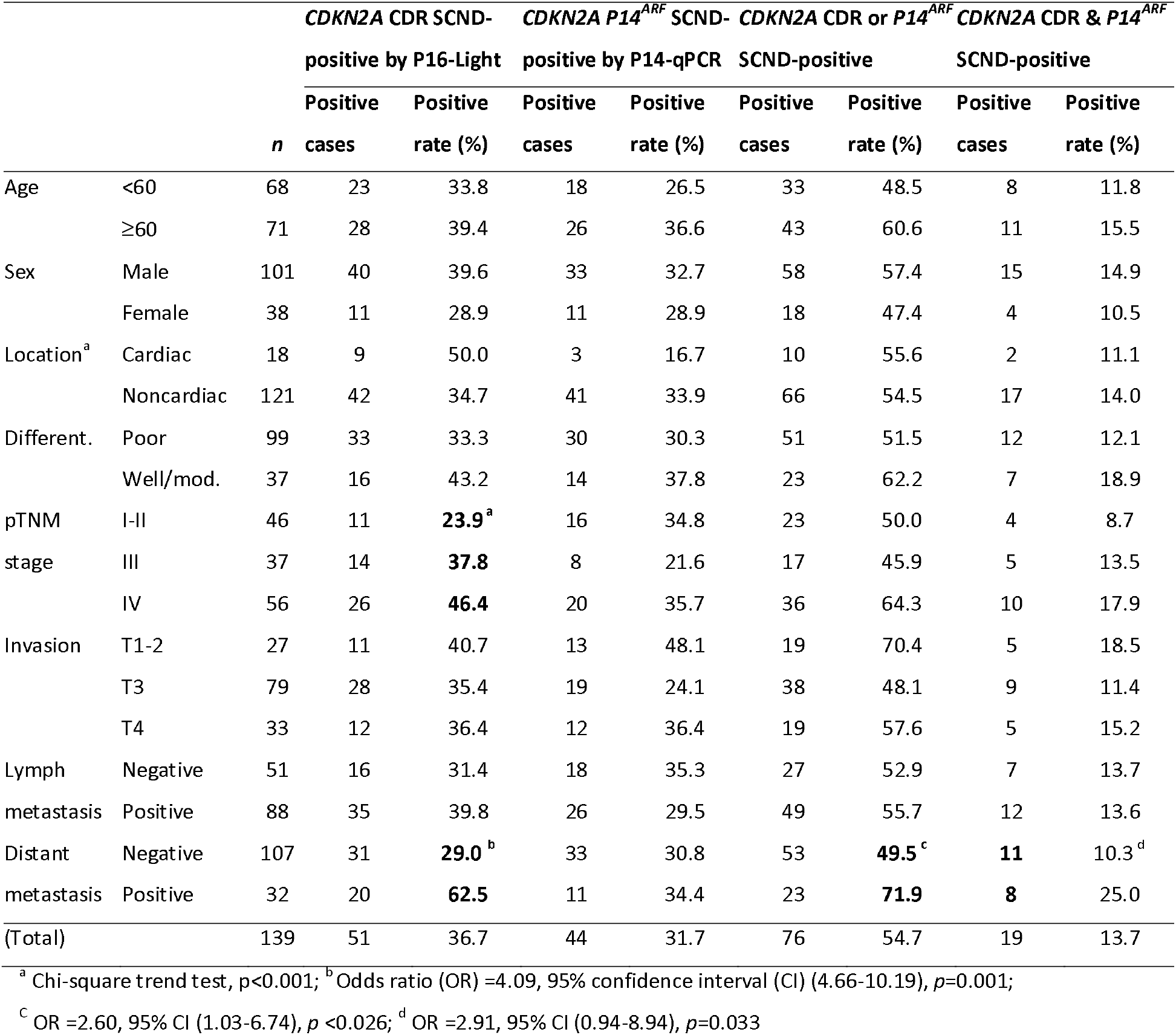
Association between clinicopathological characteristics and CDKN2A SCND detected by P16-Light and P14-qPCR

## Discussion

Somatic copy number deletion and amplification are two main kinds of SCNVs. The detection of copy number amplification of oncogenes is routinely used for precise cancer treatments. However, the detection of SCND of tumor suppressor genes is absent, and its significance in clinical practice is not well studied. The reason should be the lack of feasible detection approaches. Here, we report that there are CDRs in many tumor suppressor genes, such as *CDKN2A, miR31HG, PTEN*, and *RB1*, which are commonly inactivated by SCND in various human cancers [1,2]. Notably, we characterized, for the first time, the 5.1-kb true CDR from the *CDKN2A*/*P16*^*INK4A*^ promoter to intron-2 in >90% of cancers containing *CDKN2A* deletion. Using the *CDKN2A* CDR as a PCR target, we further established a feasible P16-Light assay to detect *CDKN2A* SCND and amplification. These findings indicate that CDRs are prevalent sequences in tumor suppressor genes, and characterization of the base-resolution genomic coordinates of CDRs could enable us to establish convenient methods for SCND detection of genes.

Interstitial deletion/fusion is the main type of *CDKN2A* SCND, and the breaking/fusing coordinates for *CDKN2A* SCNDs in cancer genomes are diverse, which blocks the establishment of a feasible detection assay for *CDKN2A* SCND, although many efforts have been made [20]. In the present study, we initially found the 8∼17-kd estimated *CDKN2A* CDR in both monoclonal cancer cell lines and cell-heterogeneous cancer tissues with *CDKN2A* copy number deletion according to the SNP-array datasets from COSMIC and TCGA projects [1,12]. Then, we further characterized the 5.1-kb true CDR at the base resolution within the *CDKN2A* gene in cancer genomes using DNA sequencing data [19-28] and confirmed the CDR using WGS datasets in all 18 GCs containing *CDKN2A* SCND [13]. Because the true *CDKN2A* CDR was observed in more than 90% of *CDKN2A*-deleted cancer samples and the P16-Light assay is highly reproducible and convenient, the quantitative P16-Light assay should be considered a viable assay for detecting *CDKN2A* SCNVs in clinical practice. This is supported by the result that *CDKN2A* SCND by P16-Light was significantly associated with GC metastasis and further supported by the results of our prospective study, in which *CDKN2A* SCND was closely associated with hematogenous metastasis of GCs [34]. In another long-term prospective study, we also found that *CDKN2A* SCND and amplification by P16-Light were significantly associated with malignant transformation and complete regression of mild or moderate esophageal squamous cell dysplasia, respectively [Fan *et al*. submitted for publication]. The results of these studies also suggest that the sensitivity of 20% for the P16-Light assay may be good enough for routine clinical use.

WGS is generally used as a kind of gold standard to study structural alterations of genomic DNAs, especially for interstitial gene copy deletion/fusions. However, WGS is a cost assay, and its accuracy depends on sequencing depth. WGS at sequencing depth 36Mendelian Inheritance in Man (OMIM) would be considered a hemi-quantitative assay. In our calling of *CDKN2A* SCND coordinate processes, it was found that 18 *CDKN2A* SCND coordinates were identified in 17 (10.8%) of 157 GCs, which was consistent with the frequency (11.4% =50/438) of homozygous deletion of *CDKN2A* in GCs in WES or WGS sequencing datasets (Data file 14) [35]. The positive rate (25.6%) of *CDKN2A* SCND in 156 GCs with enough genomic DNA samples in the P16-Light analysis was more than twice that of WGS. The results of P16-Light analyses were significantly correlated with those of WGS. These phenomena suggest that P16-Light is a much more sensitive, convenient, and less expensive assay than WGS.

P14-qPCR is a method used to detect the copy number of *CDKN2A/P14*^*ARF*^ exon-1β [15]. Although the combination of P16-Light with P14-qPCR may detect both SCNDs overlapping with the *CDKN2A* CDR and not overlapping with the *CDKN2A* CDR, the results of our comparison analysis among 139 GC patients showed that detecting *CDKN2A* SCND by individual P16-Light may be good enough for clinical practice because combination with P14-qPCR could not improve the performance of P16-Light. However, for other genes, such as *RB1* and *PTEN*, whether a qPCR array needs to be employed for detecting SCNVs should be studied case by case.

Generally, IHC is a popular method used to detect expression changes in protein-coding genes. For example, P16^INK4A^ overexpression in cervical mucosa samples is currently used for rapid HPV infection screening. We compared the status of P16^INK4A^ expression by IHC between GCs with *CDKN2A* SCND (n=4) and GCs without *CDKN2A* SCND (n=12) and did not find any difference in the P16^INK4A^ positive-staining rate between these GCs (3/4 *vs*. 9/12). The expression level of *CDKN2A/P16*^*INK4A*^ is not only affected by SCNVs but also regulated by the methylation status of CpG islands, histone modifications, and high-risk HPV infection [36,37]. These factors may partially account for the inconsistency between IHC and P16-Light.

The driver function of the *CDKN2A* gene in cancer development is enigmatic. *P16*^*ink4a*^ inactivation contributes less than *P19*^*arf*^ (the murine counterpart of human *P14*^*ARF*^) inactivation to cancer development in mice, while *P16*^*INK4A*^ inactivation contributes more than *P14*^*ARF*^ inactivation to cancer development in humans [38,39]. The exact mechanisms leading to the difference among species are still unclear. Here, we reported that approximately 87% of genetic *P16*^*INK4A*^ inactivation by *CDKN2A* SCND is accompanied by *P14*^*ARF*^ inactivation in human cancer cell lines or tissues. This may account for the species-related functional difference in the *CDKN2A* gene. The report supports this explanation that knocking out both *p16*^*ink4a*^ and *p19*^*arf*^ leads to more cancer development than individual inactivation in mice [40]. This also may account for the fact that *P14*^*ARF*^ exon-1β deletion was not associated with GC metastasis, whereas *CDKN2A* CDR deletion was significantly associated with GC metastasis, as described above.

In conclusion, we have found estimated CDRs in many tumor suppressor genes in the cancer genome. There is a 5.1-kb CDR region within the *CDKN2A* gene, and most *CDKN2A* deletions lead to *P16*^*INK4A*^ and *P14*^*ARF*^ inactivation in human cancers. Using the *CDKN2A* CDR as a target sequence, we developed a convenient quantitative multiplex PCR assay, P16-Light, to detect *CDKN2A* SCNVs in clinical practice, suggesting that the strategy to detect *CDKN2A* SCNVs may be suitable for the establishment of SCNV detection methods for other tumor suppressor genes.

## Supporting information

Supplemental Figures and Data file list

## Data Availability

Supplementary Figures 1-3 and Data files 1-14 are attached.

## Supplementary material

Supplementary Figures 1-3 and Data files 1-14 can be found online.

## Acknowledgments

We thank Dr. Sanford Dawsey at NCI, NIH, Bethesda, Maryland for critical comments on the manuscript. We also thank Miss Gina Mckeown in New York, USA for English language editing.

## Declarations

### Ethical Approval

This study was approved by the Institution Review Board of Peking University Cancer Hospital & Institute and carried out in accordance with the principles outlined in the Declaration of Helsinki. Informed consent was obtained from each patients prior to their inclusion in the study.

### Authors’ contributions

Yuan Tian: Methodology, Writing- Original draft preparation; Jing Zhou: Methodology, Formal analysis, Writing-Original draft preparation; Juanli Qiao: Investigation; Zhaojun Liu: Data curation; Liankun Gu: Investigation; Baozhen Zhang: Investigation; Youyong Lu: Supervision, Resources; Rui Xing: Conceptualization, Resources, Data curation, Validation, Writing-Original draft preparation; Dajun Deng: Conceptualization, Supervision, Funding acquisition, Methodology, Data curation, Visualization, Writing-Original draft preparation.

### Availability of data and materials

The original contributions presented in the study are included in the article as Supplemental Files 1-14.

## References

1. Beroukhim, R. et al. (2010) The landscape of somatic copy-number alteration across human cancers. Nature, 463, 899–905.

2. Cerami, E. et al. (2012) The cBio cancer genomics portal: an open platform for exploring multidimensional cancer genomics data. Cancer Discov., 2, 401–404.

3. Mermel, C.H. et al. (2011) GISTIC2.0 facilitates sensitive and confident localization of the targets of focal somatic copy-number alteration in human cancers. Genome Biol., 12, R41. doi: 10.1186/gb-2011-12-4-r41.

4. Gao, J. et al. (2013) Integrative analysis of complex cancer genomics and clinical profiles using the cBioPortal. Sci. Signal., 6, pl1. doi: 10.1126/scisignal.2004088.

5. Network CGAR. et al. (2017) Integrated genomic characterization of oesophageal carcinoma. Nature, 541, 169–175.

6. Cui, Y. et al. (2020) Whole-genome sequencing of 508 patients identifies key molecular features associated with poor prognosis in esophageal squamous cell carcinoma. Cell Res., 30, 902–913.

7. Serrano, M. et al. (1996) Role of the INK4a locus in tumor suppression and cell mortality. Cell, 85, 27–37.

8. Deng J. et al. (2018) CDK4/6 Inhibition Augments Antitumor Immunity by Enhancing T-cell Activation. Cancer Discov., 8, 216–233.

9. Jerby-Arnon, L. et al. (2018) A Cancer Cell Program Promotes T Cell Exclusion and Resistance to Checkpoint Blockade. Cell, 175, 984–997.e24.

10. Zhang, J. et al. (2018) Cyclin D-CDK4 kinase destabilizes PD-L1 via cullin 3-SPOP to control cancer immune surveillance. Nature, 553, 91–95.

11. Yu, J. et al. (2019) Genetic Aberrations in the CDK4 Pathway Are Associated with Innate Resistance to PD-1 Blockade in Chinese Patients with Non-Cutaneous Melanoma. Clin. Cancer Res., 25, 6511–6523.

12. Tate, J.G. et al. (2019) COSMIC: the Catalogue Of Somatic Mutations In Cancer. Nucl. Acids Res., 47, D941–D947.

13. Xing, R. et al. (2019) Whole-genome sequencing reveals novel tandem-duplication hotspots and a prognostic mutational signature in gastric cancer. Nat. Commun., 10, 2037. doi: 10.1038/s41467-019-09644-6.

14. Sobin, L. et al. (2009) TNM Classification of Malignant Tumours, 7th edn. International Union Against Cancer (UICC). Wiley Press, New York, USA.

15. Berggren, P. et al. (2003) Detecting homozygous deletions in the CDKN2A(p16INK4a)/ARF(p14ARF) gene in urinary bladder cancer using real-time quantitative PCR. Clin. Cancer Res., 9, 235–242.

16. Carter, S.L. et al. (2012) Absolute quantification of somatic DNA alterations in human cancer. Nat. Biotechnol., 30, 413–421.

17. Cairns, P. et al. (1995) Frequency of homozygous deletion at p16/CDKN2 in primary human tumours. Nat. Genet., 11, 210–212.

18. Novara, F. et al. (2009) Different molecular mechanisms causing 9p21 deletions in acute lymphoblastic leukemia of childhood. Hum. Genet., 126, 511–520.

19. Xie, H. et al. (2016) Mapping of deletion breakpoints at the CDKN2A locus in melanoma: detection of MTAP-ANRIL fusion transcripts. Oncotarget, 7, 16490–16504.

20. Patel, A. et al. (2014) Amplification and thrifty single-molecule sequencing of recurrent somatic structural variations. Genome Res., 24, 318–328.

21. Norris, A.L. et al. (2015) Transflip mutations produce deletions in pancreatic cancer. Genes Chromosomes Cancer 54, 472–481.

22. Guney, S. et al. (2011) Molecular characterization of 9p21 deletions shows a minimal common deleted region removing CDKN2A exon 1 and CDKN2B exon 2 in diffuse large B-cell lymphomas. Genes Chromosomes Cancer, 50, 715–725.

23. Florl, A.R. et al. (2003) Peculiar structure and location of 9p21 homozygous deletion breakpoints in human cancer cells. Genes Chromosomes Cancer, 37, 141–148.

24. Kitagawa, Y. et al. (2002) Prevalent involvement of illegitimate V(D)J recombination in chromosome 9p21 deletions in lymphoid leukemia. J. Biol. Chem., 277, 46289–46297.

25. Sasaki, S. et al. (2003) Molecular processes of chromosome 9p21 deletions in human cancers. Oncogene, 22, 3792–3798.

26. Cayuela, J.M. et al. (1997) Disruption of the multiple tumor suppressor gene MTS1/p16(INK4a)/CDKN2 by illegitimate V(D)J recombinase activity in T-cell acute lymphoblastic leukemias. Blood, 90, 3720–3726.

27. Pasmant, E. et al. (2007) Characterization of a germ-line deletion, including the entire INK4/ARF locus, in a melanoma-neural system tumor family: identification of ANRIL, an antisense noncoding RNA whose expression coclusters with ARF. Cancer Res., 67, 3963–3969.

28. Raschke, S. et al. (2005) Homozygous deletions of CDKN2A caused by alternative mechanisms in various human cancer cell lines. Genes Chromosomes Cancer, 42, 58–67.

29. Hussussian, C.J. et al. (1994) Germline p16 mutations in familial melanoma. Nat. Genet., 8, 15–21.

30. Freedberg, D.E. et al. (2008) Frequent p16-independent inactivation of p14ARF in human melanoma. J. Natl Cancer Inst., 100, 784–795.

31. Harinck, F. et al. (2012) Indication for CDKN2A-mutation analysis in familial pancreatic cancer families without melanomas. J. Med. Genet., 49, 362–365.

32. Hamosh, A. et al. (2005) Online Mendelian Inheritance in Man (OMIM), a knowledgebase of human genes and genetic disorders. Nucl. Acids Res., 33, D514–D517.

33. Amberger, J. et al. (2009) McKusick’s Online Mendelian Inheritance in Man (OMIM). Nucl. Acids Res., 37, D793–D796.

34. Qiao, J.L. et al. (2021) CDKN2A deletion leading to hematogenous metastasis of human gastric carcinoma. Frontiers in Oncology, 11, 801219. doi: 10.3389/fonc.2021.801219.

35. Liu, Y. et al. (2018) Comparative Molecular Analysis of Gastrointestinal Adenocarcinomas. Cancer Cell, 33, 721–735.e8.

36. Peters, G. (2008) Tumor suppression for ARFicionados: the relative contributions of p16INK4a and p14ARF in melanoma. J. Natl. Cancer Inst., 100, 757–759.

37. Cui, C.H. et al. (2015) P16-specific DNA methylation by engineered zinc finger methyltransferase inactivates gene transcription and promotes cancer metastasis. Genome Biol., 16, 252. doi: 10.1186/s13059-015-0819-6.

38. Li, Q. et al. (2010) Polycomb CBX7 directly controls trimethylation of histone H3 at lysine 9 at the p16 locus. PLoS One, 5, e13732. doi: 10.1371/journal.pone.0013732.

39. Li, H. et al. (2009) The Ink4/Arf locus is a barrier for iPS cell reprogramming. Nature, 460, 1136–1139.

40. Sharpless, N.E. et al. (2004) The differential impact of p16(INK4a) or p19(ARF) deficiency on cell growth and tumorigenesis. Oncogene 23, 379–385.

