## Supplemental Figures and Data file list for "Detection of Somatic Copy Number Deletion of the *CDKN2A* Gene by Quantitative Multiplex PCR for Clinical Practice"

### Supplementary Data Information

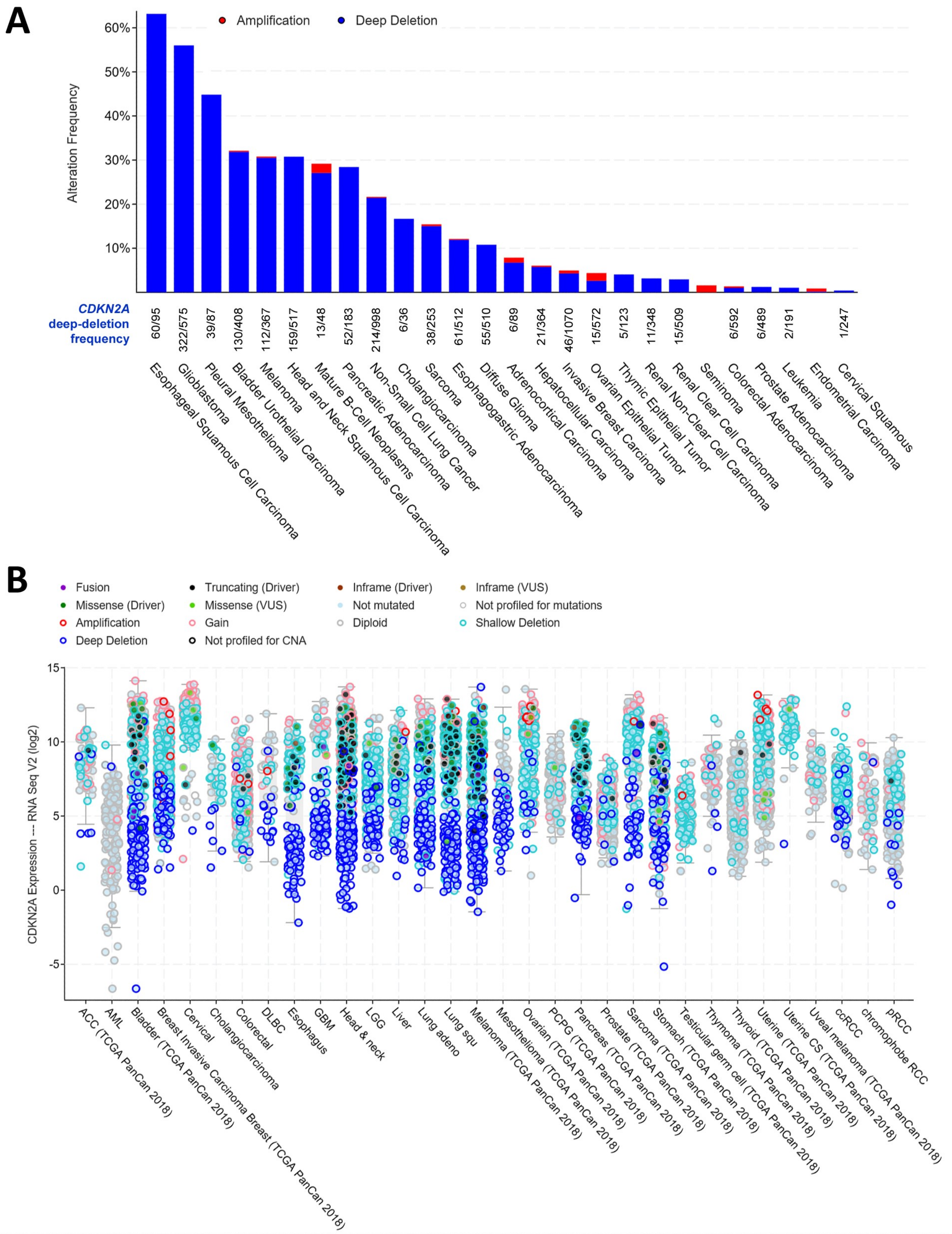


**Figure S1**. Prevalence of *CDKN2A* deep-deletion and the levels of gene expression in 10967 samples from cancer patients in Pan-TCGA studies. (**A**) Prevalence of *CDKN2A* deep deletion according to the TCGA SNP-array data. The number of total cancer cases and cases with *CDKN2A* deep-deletion are listed for each kind of cancer. (**B**) The levels of *P16$^{INK4a}$* mRNA determined by RNA sequencing in cancers with various *CDKN2A* genetic changes. The charts for patients (n=10953) in 32 Pan-TCGA studies were adapted from a graphic view at the cBioPortal Cancer Genomics website (www.cbioportal.org).

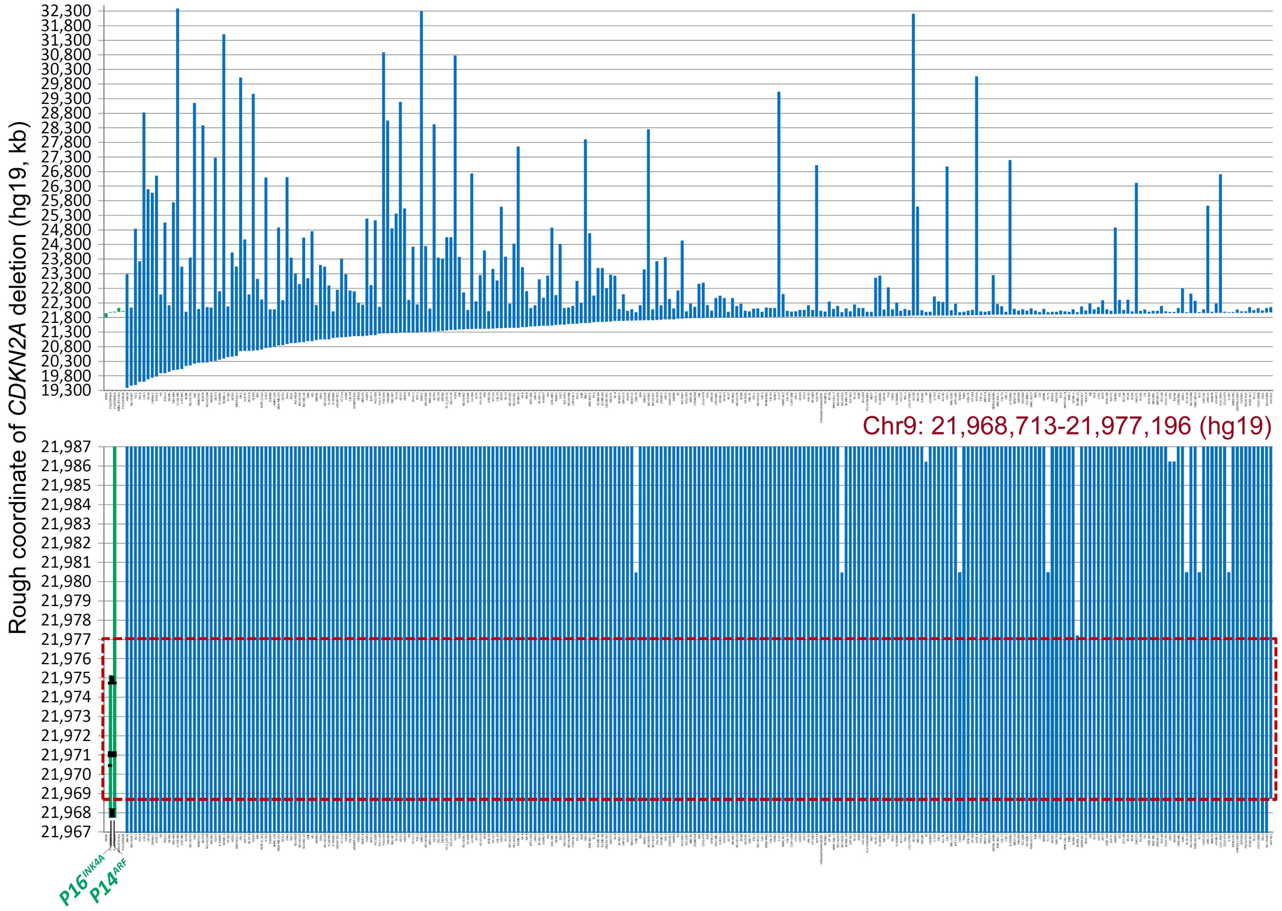


**Figure S2.** Estimated genomic coordinates of interstitial *CDKN2A* deletion/fusion in 273 human cancer cell lines with *CDKN2A* homozygous deletion according to the COSMIC data. The top chart displays the coordinates of most deletion fragments. The sample ID is labeled under each column. The bottom chart displays the amplified view of these deletion fragments, where the 8.5-kb common deletion region (CDR) is highlighted with a red dashed line rectangle. Each line represents a *CDKN2A* deletion fragment. The locations of *P16*$^{INK4a}$ and *P14*$^{ARF}$ (green lines) and exon-1α/2/3 (black dots) are also labeled as landmarks. The detailed deletion coordinates for each sample are listed in Data file 1.

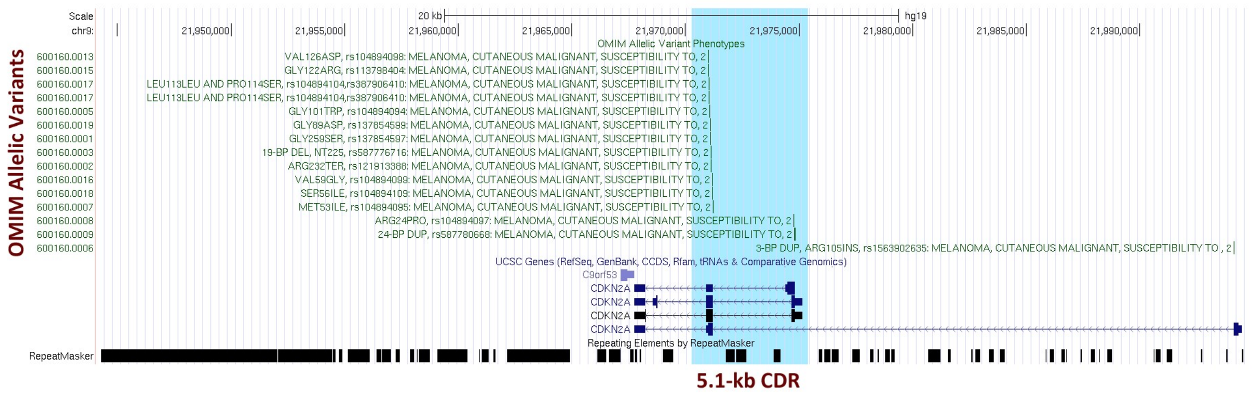


**Figure S3.** Distribution pattern of the Online Mendelian Inheritance in Man (OMIM) allelic variants within the *CDKN2A* common deletion region (CDR, highlighted in blue shadow). 12 allelic variants are located in *CDKN2A* exon-2, 2 allelic variants are located in *CDKN2A* exon-1α, and 1 allelic variant is located in *CDKN2A* exon-1β. This chart was adapted from the UCSC Genome Browser on March 10, 2021.

#### Supplementary Data file list

**Data file 1.** Estimated coordinates of *CDKN2A* deep-deletion for 273 cancer cell lines by SNP-array

**Data file 2.** Estimated coordinates of *ATM* deep-deletion for TCGA cancers by SNP-array

**Data file 3.** Estimated coordinates of *CDKN2A* deep-deletion for TCGA cancers by SNP-array

**Data file 4.** Estimated coordinates of *FAT1* deep-deletion for TCGA cancers by SNP-array

**Data file 5.** Estimated coordinates of *miR31HG* deep-deletion for TCGA cancers by SNP-array

**Data file 6.** Estimated coordinates of *PTEN* deep-deletion for TCGA cancers by SNP-array

**Data file 7.** Estimated coordinates of *RB1* deep-deletion for TCGA cancers by SNP-array

**Data file 8.** Estimated coordinates of *CCSER1* deep-deletion for TCGA cancers by SNP-array

**Data file 9.** Estimated coordinates of *FHIT* deep-deletion for TCGA cancers by SNP-array

**Data file 10.** Estimated coordinates of *LRP1B* deep-deletion for TCGA cancers by SNP-array

**Data file 11.** Estimated coordinates of *WWOX* deep-deletion for TCGA cancers by SNP-array

**Data file 12.** True coordinates of *CDKN2A* interstitial deletion/fusion for 110 cancer cell lines and tissues by sequencing

**Data file 13.** The status for *CDKN2A* SCNV in gastric cancer samples and sample purity

**Data file 14.** *CDKN2A* homozygous deletion in STADs from patients by WGS or WES
